# Obstetric Complications Associated With Adolescent Pregnancy At Livingstone University Teaching Hospital

**DOI:** 10.1101/2025.06.01.25328749

**Authors:** Natasha Chishala, Joreen P. Povia, Nestorine N. Ngongo, Joel M. Chisanga, Timothy Mabuku, Emmanuel Yumba, Emmanuel Riwo, Salma M. Baines, Prince Mulambo, Steven Kumeleni, Lukundo Siame, Bislom C. Mweene, Katongo H. Mutengo, Chileleko Siakabanze, Emmanuel Luwaya, Hanzooma Hatwiko, Patson Sichamba, Martin Chakulya, Sepiso K. Masenga

**Affiliations:** Department of Pathology, Mulungushi University, School of Medicine and Health Sciences, Livingstone, Zambia; Department of Health Economics, Division of Integrated Sciences, Livingstone Center for Prevention and Translational Science, Livingstone, Zambia; Department of Medicine, Division of Integrated Sciences, Livingstone Center for Prevention and Translational Science, Livingstone, Zambia

**Keywords:** adolescent pregnancy, gestational age, maternal morbidity, Zambia, resource-limited settings, obstetric complications, preterm labour, preeclampsia

## Abstract

**Background:** Adolescent pregnancy remains a major public health concern in sub-Saharan Africa, where limited data exist on region-specific obstetric complications. This study aimed to identify factors associated with maternal morbidity among adolescents at a tertiary referral hospital in Zambia.

**Methods:** A retrospective cross-sectional study was conducted using records of 407 adolescents (10– 19 years) admitted to Livingstone University Teaching Hospital (LUTH) between January 2023 and December 2024. Sociodemographic, clinical, and laboratory variables were evaluated against primary outcomes (preterm labour, preeclampsia/eclampsia, haemorrhage) using multivariable logistic regression. Data were analysed in Statcrunch, with results reported as adjusted odds ratios (AORs) and 95% confidence intervals (CIs).

**Results:** The median age was 17 years (IQR 16–18); 90 adolescents (22.1%) experienced ≥1 maternal complication. In multivariable analysis, adverse birth outcomes (preterm delivery or intrauterine death) were associated with 14-fold higher odds of maternal complications (adjusted odds ratio [AOR] 14.3, 95% CI 4.9–42.1; p < 0.0001). Each additional week of gestation was protective (AOR 0.7, 95% CI 0.6–0.8; p < 0.0001). Other factors including age, residence, employment status, parity, foetal complications, and proteinuria were not independently significant.

**Conclusions:** Nearly one in five adolescent pregnancies at LUTH is complicated by serious maternal morbidity. Adverse birth outcomes and gestational age emerge as powerful independent predictors. These findings underscore the urgent need for interventions to prevent pre-eclampsia, optimize timing of delivery, and improve perinatal care among adolescents in resource-constrained settings. Prospective, multicentre studies including neonatal outcomes are warranted to guide comprehensive adolescent-focused maternal health strategies.

## Introduction

Adolescent pregnancy remains a significant public health challenge, particularly in low- and middle-income countries (LMICs), where limited access to education, healthcare, and contraception perpetuates high rates of early childbearing [1,2]. Globally, approximately 21 million girls aged 15–19 years in developing regions become pregnant annually, with complications during pregnancy and childbirth representing the leading cause of mortality in this age group [3,4]. In sub-Saharan Africa (SSA), the adolescent fertility rate is the highest worldwide, averaging 101 births per 1,000 girls aged 15–19, with Zambia reporting a rate of 132 births per 1,000 adolescents nearly double the global average [1,5]. These pregnancies are fraught with obstetric risks, including preterm birth, low birth weight, preeclampsia, and obstructed labour, driven by biological immaturity and socioeconomic vulnerabilities such as poverty, early marriage, and inadequate prenatal care [6,7].

In Zambia, 28% of women aged 15–19 years have begun childbearing, with rural adolescents disproportionately affected [8]. At Livingstone University Teaching Hospital (LUTH), a tertiary referral centre in Southern Province of Zambia, adolescent mothers constitute 18–25% of annual deliveries, reflecting national trends [9,10]. Preliminary data from LUTH’s maternity ward highlight alarming rates of complications: 35% of adolescent pregnancies are complicated by anaemia, 22% by preterm delivery, and 15% by hypertensive disorders, compared to 10–15% in adults [11]. Furthermore, perinatal mortality among adolescents at LUTH is reported at 12%, nearly twice the rate observed in older mothers [9]. These disparities underscore the urgent need to contextualize the burden of adolescent pregnancy and its obstetric outcomes in resource-constrained settings [10,12].

Over 60% of pregnancies in adolescents are unintended, exacerbated by limited sexual health education and cultural norms prioritizing early marriage [12,13]. Health system challenges, including delayed antenatal care initiation and shortages of skilled birth attendants, further exacerbate risks [11]. This study aimed to quantify the prevalence and spectrum of obstetric complications associated with adolescent pregnancy at LUTH, providing evidence to guide targeted interventions and mitigate the dual burden of maternal and neonatal morbidity in Southern Province of Zambia.

## Methodology

### Study Design

This retrospective cross-sectional study utilized hospital records to evaluate serious maternal complications associated with adolescent pregnancy (ages 10–19 years) at Livingstone University Teaching Hospital (LUTH), Zambia.

### Study setting

The study was conducted at LUTH, a tertiary referral hospital in Southern Province, Zambia, serving a high-risk, high-volume obstetric population. The facility’s extensive catchment area and annual delivery load provided a robust sample for assessing adolescent pregnancy complications.

### Eligibility and Recruitment

From a total of 1200 available files, we screened 820 files for adolescent pregnancies who delivered or were admitted to the hospital between the period of January 2023 and December 2024, aging between 10 to 19years. A total of 407 files were eligible and analysed. A total of 413 files were excluded due to absence of predefined complications (e.g., preterm labour, pre-eclampsia, postpartum haemorrhage) and missing data of interest including age, sex, residence, other relevant maternal details and diagnosis.

### Data Collection

A structured retrospective review was conducted, and we accessed and abstracted the data starting on 2^nd^ January 2025 and completed the abstraction on 16^th^ April, 2025, by the aid of trained research assistants using a standardized protocol. Data abstraction was performed via the Research Electronic Data Capture (REDCap) platform to ensure consistency and minimize entry errors.

### Study Variables

The primary outcome variable was maternal complications and was defined by the presence of preterm labour, pre-eclampsia, eclampsia, postpartum haemorrhage, and antepartum haemorrhage. Maternal complications were defined using WHO criteria: preterm labour (regular uterine contractions with cervical changes before 37 weeks) [6], pre-eclampsia (new hypertension ≥140/90 mmHg after 20 weeks with proteinuria ≥1+ dipstick or end-organ dysfunction), eclampsia (tonic-clonic seizures in pre-eclampsia excluding other causes) [14], postpartum haemorrhage (blood loss ≥500 mL vaginal/≥1000 mL caesarean or clinical instability), and antepartum haemorrhage (vaginal bleeding ≥20 weeks, excluding trauma).

Independent Variables included *Sociodemographic*: Age, residence (urban/rural), employment status, education level. *Clinical*: Parity, gravidity, delivery mode, adverse birth outcomes (based on either preterm birth or death), blood pressure, proteinuria, serum creatinine, ALT, haematological indices (WBC, RBC, haemoglobin, platelets), and serum urea.

### Data analysis

We exported data from the REDCap application to Microsoft Excel 2013, where it underwent cleaning and coding. Statcrunch was then used for data analysis. Descriptive statistics were employed to summarize categorical variables through frequencies and percentages, while continuous variables were summarized using the median and interquartile range. The Shapiro-Wilk test was applied to evaluate data normality. The association between two categorical variables was assessed using the chi-square test, and differences between two medians were evaluated with the Wilcoxon rank-sum test. Univariable and multivariable logistic regression analyses were conducted to identify factors associated with obstetrics complications.

### Ethical considerations

Ethical approval was granted by the Mulungushi University School of Medicine and Health Sciences Research Ethics Committee (Ref: SMHS-MU2-2024-238) on May 10, 2024. All data were de-identified to protect confidentiality, and no personally identifiable information was collected. As the study utilized retrospective, anonymized records, informed consent was waived by the ethics committee.

The Strengthening the Reporting of Observational Studies in Epidemiology (STROBE) guidelines were followed to ensure methodological rigor, Supplementary file 1.

## Results

From 1200 available files for abstraction, we reviewed 820 files. A total of 407 were analysed, while 413 were excluded due to absence of age and predefined complications (e.g., preterm labour, pre-eclampsia, postpartum haemorrhage), Fig 1.

**Figure 1.**
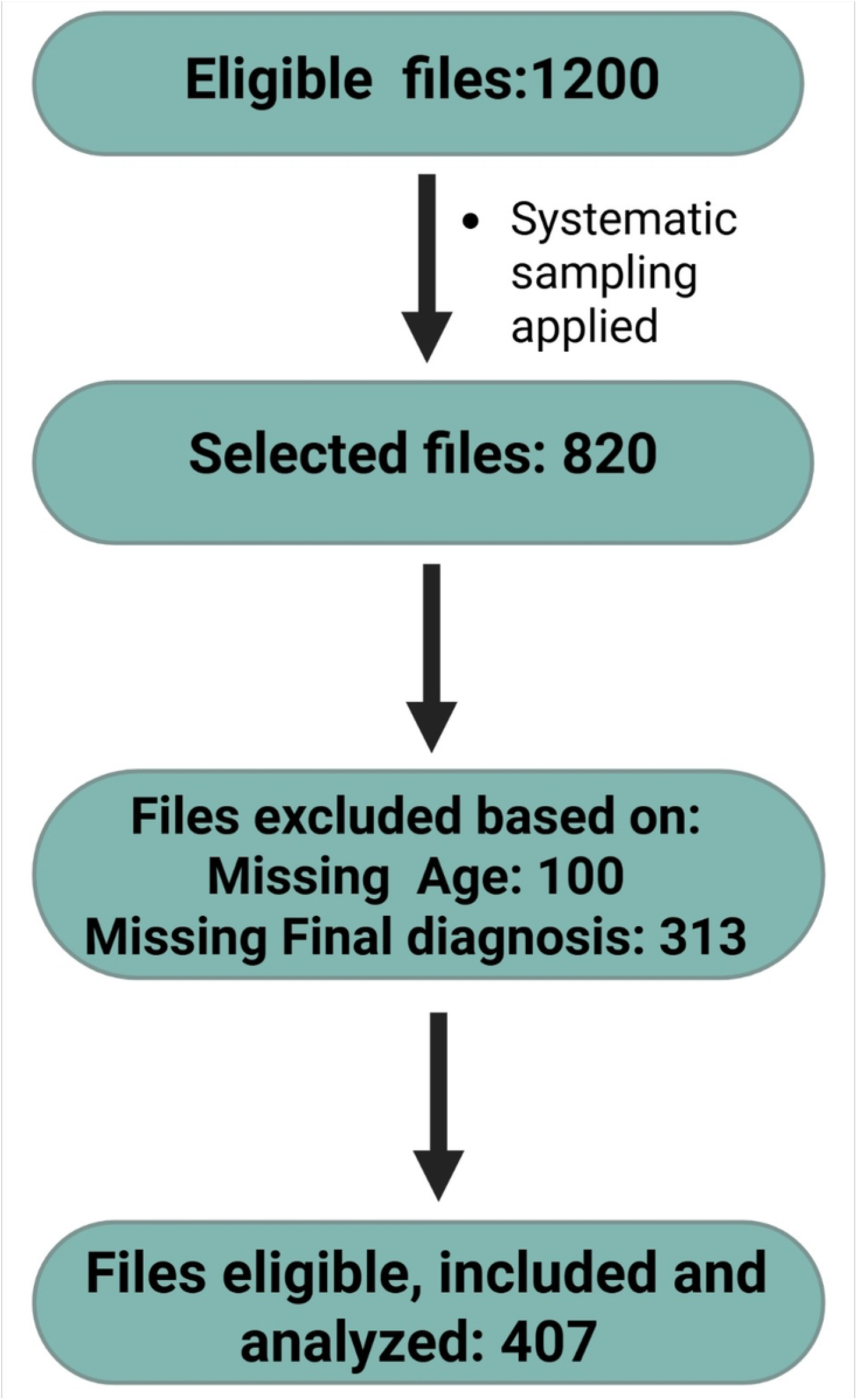
Eligibility flow diagram

### Characteristics of the study population

The study included 407 adolescent participants with maternal complications, with a median age of 17 years (IQR: 16–18) (Table1). No significant age difference was observed between those with complications (18 years, IQR: 16–18) and those without (17 years, IQR: 16–18; p = 0.153). Most participants were unemployed 97.1% (n=395), resided in rural areas 97.1% (n=395), and had primary education 59.7% (n=243). Majority of birth outcomes were term births 84.5% (n=344), while preterm births were more 12.3% (n=50) than deaths 3.2% (n=13). Most adolescent pregnant mothers didn’t have any history of pre-eclampsia 99.8% (n=406), current pre-eclampsia 94.5% (n=385) and eclampsia 97.5% (n=397). This is in association with proteinuria on dipstick with 96.6% (n=393).

**Table 2. Factors associated with adolescent pregnancy complications in Logistic Regression.**

In the univariable analysis, birth outcomes was strongly associated with maternal complications, with 29.4 times higher odds (95% CI: 14.1–61.2, p < 0.0001), Table 2. Those who reported to currently have preeclampsia had a very large effect size (OR = 27.9, 95% CI: 8.0–97.0, p < 0.0001), while gestational age showed protective effect, with each unit increase associated with 30% lower odds of the outcome (OR = 0.7, 95% CI: 0.6–0.8, p < 0.0001). Additionally, Foetal complication was significantly association with the outcome (OR = 3.2, 95% CI: 2.2–4.6, p < 0.0001). Proteinuria on dipstick had strong association with the outcome (OR = 9.7, 95% CI: 3.0–31.9, p = 0.0002). In multivariable analysis, birth outcomes had attenuated magnitude but retained strong significance (AOR = 14.3, 95% CI: 4.9–42.1, p < 0.0001) while those who reported to currently have preeclampsia showed a dramatic effect size increase (AOR = 44.7, 95% CI: 7.3–275.2, p < 0.0001), and gestational age still showed protective effect (AOR = 0.7, 95% CI: 0.6–0.8, p < 0.0001).

**Table 1.**
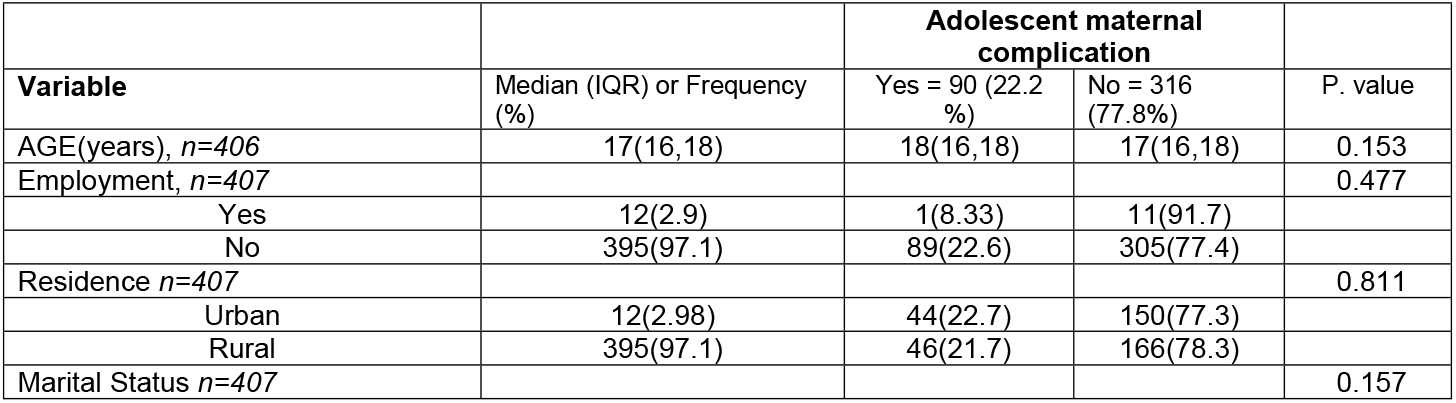
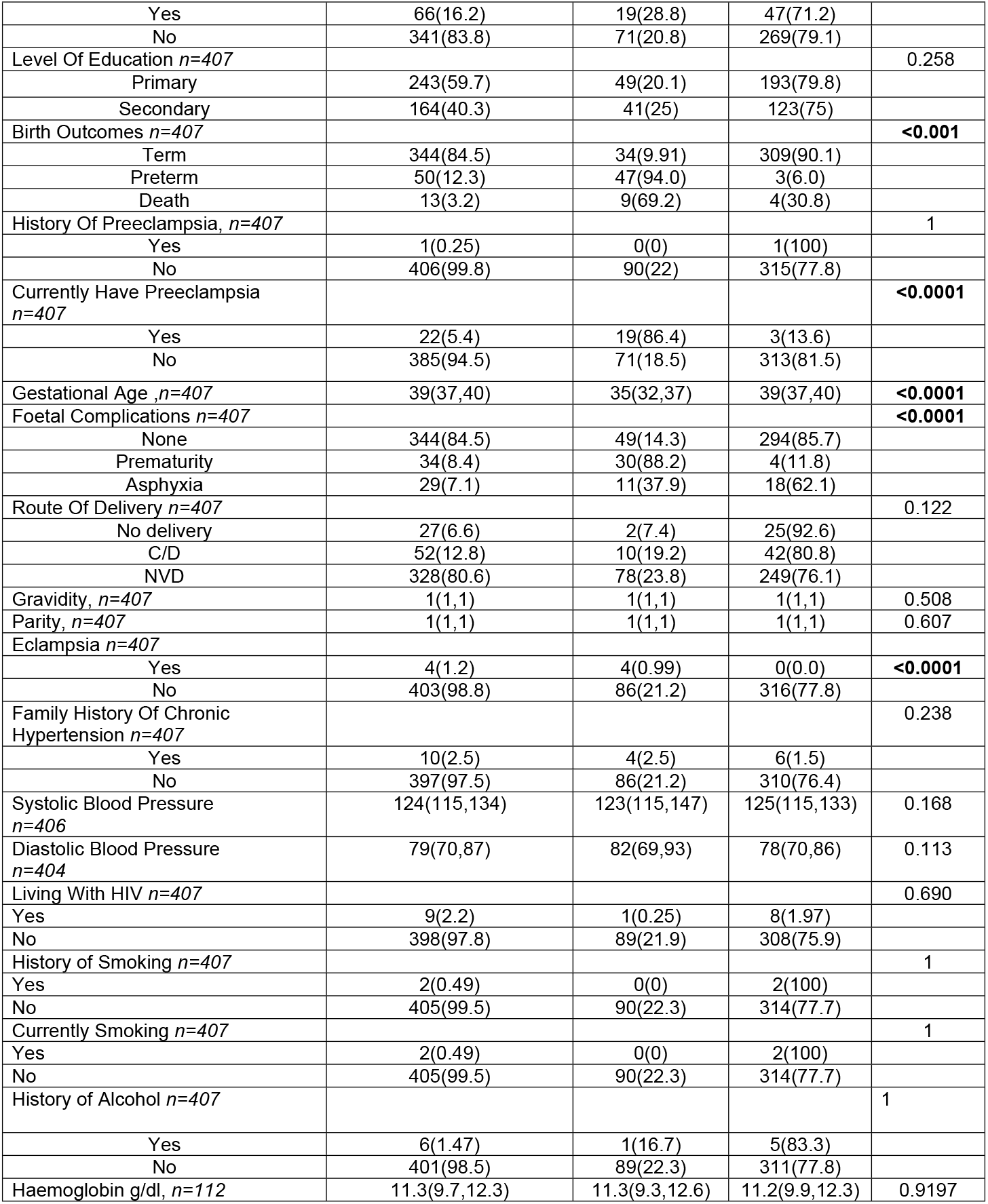
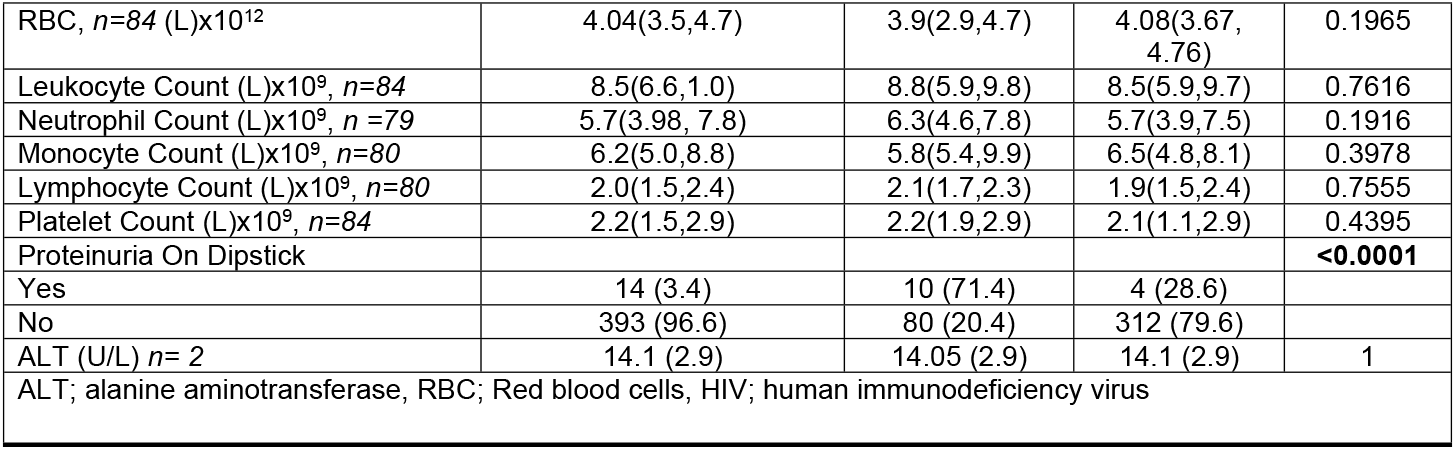
Clinical and demographic characteristics associated with adolescent pregnancy complications

**Table 2.** Factors associated with Adolescent maternal complication

| Table 2. Factors associated with Adolescent maternal complication |  |  |  |  |
| --- | --- | --- | --- | --- |
|  | Univariable |  | Multivariable |  |
| Variables | OR (95%CI) | p. value | AOR (95%CI) | p. value |
| **Age** | 1.1(0.9,1.3) | 0.1664 | 1.0(0.7,1.3) | 0.787 |
| **Adverse Birth outcomes** | 29.4(14.1,61.2) | **<0.0001** | 14.3(4.9,42.1) | **<0.0001** |
| **Gestational age** | 0.7(0.6,0.8) | **<0.0001** | 0.7(0.6,0.8) | **<0.0001** |
| **Fetal complications** | 3.2(2.2,4.6) | **<0.0001** | 1.2(0.6,2.4) | 0.6945 |
| **Proteinuria on dipstick** | 9.7(3.0,31.9) | **0.0002** | 2.5(0.2,26.7) | 0.448 |
| Abbreviations: OR; odds ratio, AOR; adjusted odds ratio |  |  |  |  |

## Discussion

Our study at the Livingstone University Teaching Hospital (LUTH) investigated obstetric complications in adolescent pregnancies. Adverse birth outcomes (preterm birth or death) emerged as a strong independent marker of risk. This finding aligns with registry data indicating that complications such as preterm birth and growth restriction not only heighten immediate perinatal danger but also portend longer-term cardiovascular risks for mothers and offspring [15]. A study by [16] supports this evidence and highlight the increased incidence of adverse birth outcomes in teenage pregnancies and their potential sequelae. Similarly, research across Africa has consistently demonstrated this vulnerability and revealed a significant association between adolescent pregnancies and a higher likelihood of preterm birth, low birth weight, and stillbirth [17]. These findings resonate with studies conducted elsewhere, such as the one in Sweden where women experiencing multiple pregnancy complications had markedly increased rates of ischemic heart disease later in life [18].Furthermore, large-scale analyses have linked hypertensive pregnancies to elevated rates of neonatal complications, including metabolic disturbances, which further highlights the downstream impact of adverse birth events [19].

Gestational age was significant to the outcome despite the odds ratio indicating a protective effect against adverse maternal-foetal outcomes, these findings should be interpreted cautiously due to the study’s limited sample size. The small sample reduces statistical power, increases vulnerability to chance effects, and may inflate the magnitude of observed associations, underscoring the need for validation in larger, more robust cohorts. However, the results aligns with large, population-based cohorts demonstrating that adolescent pregnancy complications greatly increase preterm birth risk, and that each additional week in utero meaningfully lowers morbidity [25,26]. Our findings are in agreement with [27] which reported substantially higher odds of preterm delivery among women with hypertensive disorders compared to normotensive pregnancies. Collectively, these data underscore that even modest prolongation of pregnancy can yield significant health benefits.

## Strengths

This study possesses several notable strengths that contribute to its significance. Directly our study addresses the critical public health issue of adolescent pregnancy and its associated obstetric complications, a significant concern particularly in low- and middle-income countries. By focusing on this vulnerable population, the research provides valuable data relevant to improving their health outcomes. The study’s setting at Livingstone University Teaching Hospital (LUTH), a tertiary referral centre in Zambia with a high adolescent fertility rate, ensures that the findings are contextually relevant to similar resource-constrained environments. The high volume of obstetric cases at LUTH also provided a substantial sample size for analysis. Furthermore, the study’s methodological rigor is a key strength. The use of standardized WHO criteria for defining obstetric complications enhances the internal validity of the findings and allows for better comparison with international research. The retrospective cross-sectional design, while having limitations, was an efficient approach to assess the prevalence of complications and explore associations using existing hospital records. The application of appropriate statistical methods, including descriptive statistics and logistic regression, enabled the identification of significant relationships between various factors and maternal complications. Notably, the study successfully identified adverse birth outcomes and current preeclampsia as strong independent predictors of maternal complications within this adolescent cohort. Finally, the study’s adherence to ethical guidelines, including obtaining ethical approval and ensuring data confidentiality through de-identification, is commendable. The use of REDCap for data collection likely contributed to improved data accuracy and management.

## Limitations

Despite its strengths, the study also has several limitations that need to be considered when interpreting the findings. The retrospective design, relying on existing hospital records, introduces potential biases related to data completeness and accuracy. The exclusion of a significant proportion of files due to missing data on crucial variables raises concerns about potential selection bias and the generalizability of the results to the entire population of adolescent pregnancies at LUTH. The cross-sectional nature of the study also limits the ability to establish temporal relationships or causality between the identified risk factors and maternal complications. The findings demonstrate associations but cannot definitively prove that these factors directly caused the complications. Another limitation is the single-centre design, which may restrict the generalizability of the findings to other healthcare facilities in Zambia, particularly those at primary and secondary care levels, and to adolescents in different geographical regions with varying socioeconomic conditions and access to healthcare. The study’s exclusive focus on maternal complications, while providing valuable insights, omits the crucial aspect of neonatal outcomes, limiting a comprehensive understanding of the overall impact of adolescent pregnancy. The statistically significant inverse association observed between gestational age and maternal complications, along with its relatively narrow confidence interval, requires cautious interpretation in light of the study’s definition of complications, which included preterm labour, preeclampsia, eclampsia antepartum haemorrhage and postpartum haemorrhage. Finally, the limited sample size for certain sub-analyses, such as those involving specific medical histories or laboratory parameters, restricts the statistical power to detect significant associations for these variables, and the potential for unmeasured confounding factors inherent in retrospective observational studies cannot be entirely ruled out.

## Conclusion

This retrospective cross-sectional study, conducted at a major tertiary referral hospital in Zambia, provides compelling evidence of the substantial burden of obstetric complications among adolescent pregnancies. Our analysis robustly identifies adverse birth outcomes and the presence of current preeclampsia as potent and independent predictors of maternal complications within this vulnerable population. We found strikingly (a 14.3-fold) increased risk of maternal complications in adolescents experiencing adverse birth outcomes. While the statistically significant inverse association observed with gestational age requires nuanced interpretation in the context of our composite outcome definition, it highlights the inherent risks associated with earlier gestational ages and the importance of preventing preterm births.

Despite the valuable insights gained, the inherent limitations of our retrospective, single-centre design necessitate cautious extrapolation of these findings to the broader population of pregnant adolescents in Zambia. The potential for selection bias due to missing data and the cross-sectional nature of the study, which precludes the establishment of causality, warrant further investigation through well-designed prospective, multi-centre studies with robust sample sizes. Future research should also prioritize the inclusion of neonatal outcomes to provide a more comprehensive understanding of the intergenerational impact of adolescent pregnancy and explore the complex interplay of socio-cultural determinants and healthcare access in shaping these adverse outcomes. Nevertheless, our findings serve as a critical call to action for policymakers, healthcare providers, and public health stakeholders in Zambia and similar LMICs. Targeted interventions focusing on preventing unintended adolescent pregnancies, improving access to quality antenatal care with early identification and management of preeclampsia, and addressing the underlying socioeconomic vulnerabilities are essential to mitigate the disproportionate burden of obstetric complications and ultimately improve maternal and child health outcomes in this high-risk population.

## Data Availability

The raw data underlying the results presented in the study have been uploaded as supporting information.

## Author’s contributions

NC and SKM conceived the Study. SKM and NC oversaw Data acquisition. SKM, MC, and NC supervised data acquisition. SKM and NC conducted the formal analysis. NC, JPP, NNN, JMC, TM, EY, WR, SMB, PM, SK, LS, BCM, KHM, CS, EL, HH, MC and SKM wrote the original draft. All authors contributed to the article edits and approved the final manuscript.

## Supplementary files

S1. Strobe checklist

S2. Data

